# An atlas of associations between polygenic risk scores from across the human phenome and circulating metabolic biomarkers

**DOI:** 10.1101/2021.10.14.21265005

**Authors:** Si Fang, Michael V Holmes, Tom R Gaunt, George Davey Smith, Tom G Richardson

## Abstract

Polygenic risk scores (PRS) are becoming an increasingly popular approach to predict complex disease risk, although they also hold the potential to develop insight into the molecular profiles of patients with an elevated genetic predisposition to disease. In this study, we have constructed an atlas of associations between 129 different PRS and 249 circulating metabolites in up to 83,004 participants from the UK Biobank study. As an exemplar to demonstrate the value of this atlas we conducted a hypothesis-free evaluation of all associations with glycoprotein acetyls (GlycA), an inflammatory biomarker. Using bi-directional Mendelian randomization, we find that the associations highlighted likely reflect the effect of risk factors, such as body mass index (Beta=0.16 per standard deviation change in GlycA, 95% CI=0.11 to 0.21, P=9.9×10^−10^) or liability towards smoking cigarettes (Beta=0.28, 95% CI=0.20 to 0.35, P=2.4×10^−14^), on systemic inflammation as opposed to the converse direction of effect. Furthermore, we repeated all analyses in our atlas within age strata to investigate potential sources of collider bias, such as medication usage. This was exemplified by comparing associations between lipoprotein lipid profiles and the coronary artery disease PRS in the youngest and oldest age strata, which had differing proportions of individuals undergoing statin therapy. All results can be visualised at http://mrcieu.mrsoftware.org/metabolites_PRS_atlas.

## Introduction

Complex traits and disease have a polygenic architecture meaning that they are influenced by many genetic variants scattered throughout the human genome (Boyle *et al*., 2017). An increasingly popular approach to predict disease risk in a population is to derive weighted scores by summing the number of risk increasing variants that participants harbour. These are typically referred to as ‘polygenic risk scores’ (PRS) (Torkamani *et al*., 2018, Lewis and Vassos, 2020). In the last decade, PRS have emerged as powerful tools for predicting lifelong risk of disease, which is predominantly due to the dramatic increase in sample sizes of genome-wide association studies (GWAS) and their continued success in uncovering trait-associated genetic variants across the genome (Visscher *et al*., 2017). Additionally, PRS have utility in a causal inference setting to establish causal effects between risk factors and disease outcomes, as well as to help elucidate putative diagnostic and prognostic biomarkers for disease incidence (Richardson *et al*., 2019b, Holmes and Davey Smith, 2019).

The human metabolome consists of over 100,000 small molecules and is a rich source of potential risk factors and biomarkers, as well as therapeutic targets (Holmes *et al*., 2021), for complex traits and disease (Gallois *et al*., 2019). Many circulating metabolic traits studied to date have a large heritable component as demonstrated by GWAS endeavours (Suhre *et al*., 2011, Shin *et al*., 2014, Lotta *et al*., 2021), suggesting that they have a polygenic architecture. In-depth molecular profiling has recently been undertaken in the UK Biobank (UKB) study using nuclear magnetic resonance (NMR) to capture measures of 249 circulating metabolites in approximately 120,00 participants who also have genotype data (Julkunen *et al*., 2021, Sudlow *et al*., 2015). This resource therefore provides an unprecedented opportunity to characterize metabolic profiles for disease risk by leveraging genome-wide variation captured by PRS. There are multiple advantages to this approach over conventional observational associations between metabolites and complex traits or endpoints. For example, as UKB is a prospective cohort study many diseases have low prevalence, such as Alzheimer’s disease which typically has a late onset. In contrast, evaluations using PRS will likely yield higher statistical power given that a continuous genetic score will be analysed for all participants in UKB based on their liability to disease.

In this study, we sought to construct an atlas of associations between 129 PRS and the 249 circulating metabolic traits in the UKB study (**Figure 1**). We demonstrate the usefulness of this atlas in terms of highlighting putative risk factors and biomarkers for disease risk and advocate the use of an approach known as Mendelian randomization (MR) to investigate whether a causal relationship may underlie findings (**Supplementary Note 2**) (Davey Smith and Ebrahim, 2003, Davey Smith and Hemani, 2014). As an exemplar, we apply MR systematically to investigate all PRS associations highlighted from a hypothesis-free scan of the inflammatory marker glycoprotein acetyls. Furthermore, all PRS analyses were initially conducted in the full UKB sample, as well as in age tertiles as proposed previously to evaluate the influence of medication use on findings (Bell *et al*., 2021). As the age of individuals in UKB is very unlikely to induce sources of biases into analyses (e.g. collider bias), age-dependent stratification allows comparisons between the youngest and oldest tertiles in UKB where the level of medication use is likely to vary between groups. Our findings provide valuable insights into the effects of PRS on metabolic markers which may influence hypothesis generation and facilitate similar analyses to those presented in this paper.

**Figure 1:**
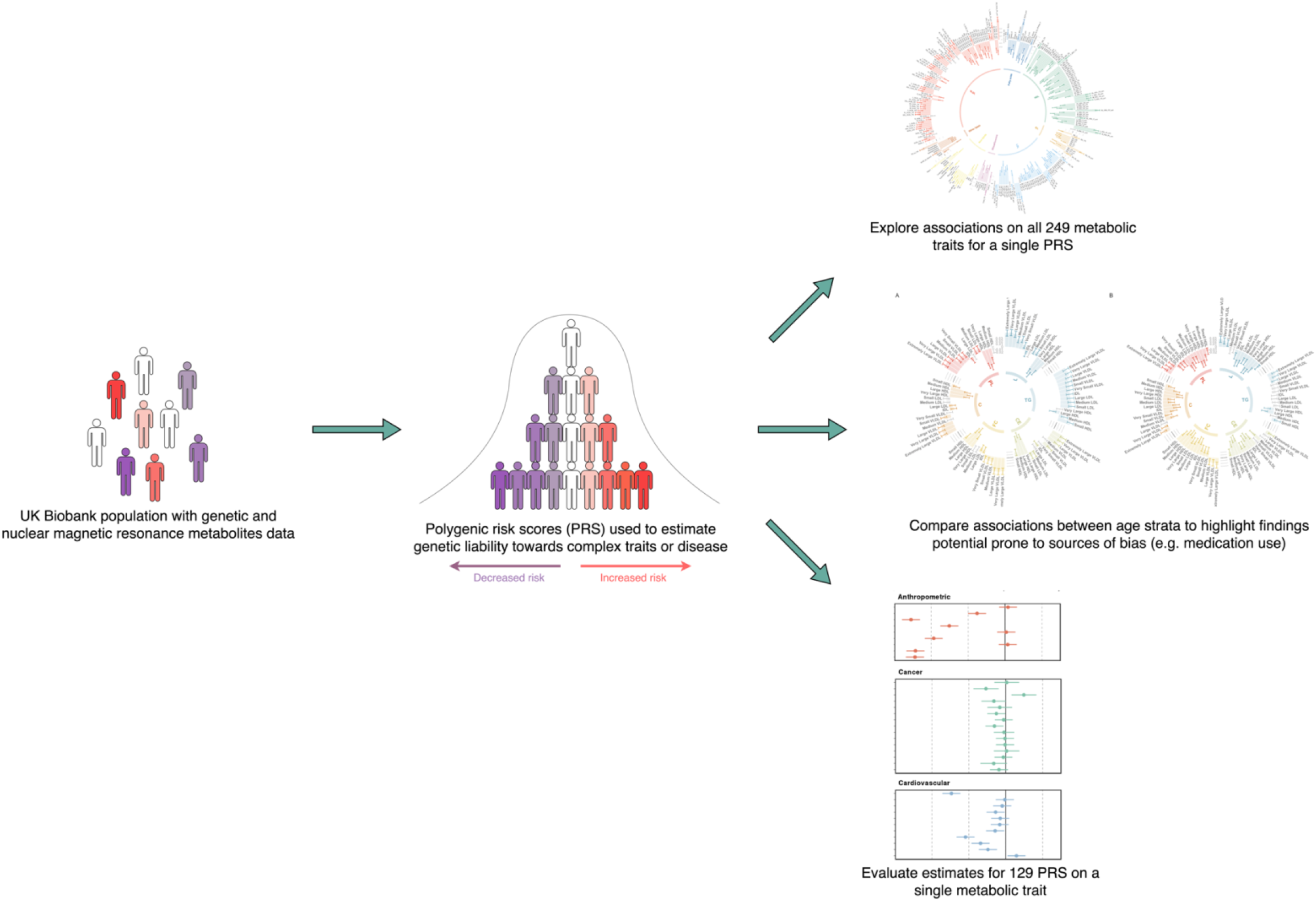
A schematic diagram depicting the analytical approach undertaken in this study

## Results

### Constructing an atlas of polygenic risk score associations across the human metabolome

We obtained genome-wide summary statistics for 129 different complex traits and diseases from large-scale GWAS and constructed PRS for each of these in the UKB study. The majority of these summary statistics were obtained from the OpenGWAS platform and encompassed traits and disease outcomes from across the human phenome (Elsworth *et al*., 2020). GWAS were identified based on those conducted in populations of European descent given that our analysis in UKB was based on the European subset with NMR data. Furthermore, we identified studies which did not include the UKB in their study to avoid overlapping samples between PRS construction and analysis with metabolic traits. Full details for all GWAS can be found in **Supplementary Table 1**. Two versions of each PRS were built using different thresholds for variant-trait associations (P) and linkage disequilibrium (LD; r^2^). These were 1) “lenient” thresholds of P<0.05 & r^2^<0.1 and 2) a “stringent” threshold of P<5×10^−8^ and r^2^<0.001. PRS were generated using the software PLINK (Chang *et al*., 2015) with LD being calculated using a reference panel of 10,000 randomly selected unrelated UKB individuals of European descent (Kibinge *et al*., 2020).

We investigated the association between each PRS in turn with 249 circulating metabolites measured using targeted high-throughput NMR metabolomics from Nightingale Health Ltd (biomarker quantification version 2020) (**Supplementary Table 2**)(Julkunen *et al*., 2021). Amongst these traits were biomarkers quantifying lipoprotein lipid traits, their concentrations and subclasses, fatty acids, ketone bodies, glycolysis metabolites and amino acids. Our final sample size of n=83,004 was determined based on individuals with both genotype and circulating metabolites data after removing participants with withdrawn consent, evidence of genetic relatedness or who were not of ‘white European ancestry’ based on a K-means clustering (K=4). All PRS were standardized to have a mean of 0 and standard deviation of 1 and similarly all metabolites were subject to inverse rank normalization transformations prior to analysis allowing cross-PRS/metabolite comparisons to be made. Analyses were conducted using linear regression adjusting for age, sex and the top 10 principal components.

To disseminate all findings from this large-scale analysis we have developed a web application (http://mrcieu.mrsoftware.org/metabolites_PRS_atlas/) to query and visualise metabolic signatures for a given PRS. In this paper, we have discussed findings using PRS that were derived using the more lenient criteria (i.e., P<0.05 & r^2^<0.1), although all findings based on both thresholds can be found in the web atlas. In total, there were 5,721 associations between PRS derived using this criteria and NMR-assessed circulating metabolic traits (based on false discovery rate (FDR) <5% in each PRS analysis). Heatmaps depicting all PRS-metabolic trait associations can be found in **Supplementary Figures 1 & 2**. The PRS with the largest number of associations robust to FDR corrections was body mass index (BMI) (n=226) (**Supplementary Table 3**).

Many top associations across PRS were consistent with the known underlying biology of the disease used to weight these scores, as well as various proof of concepts that PRS are capable of predicting their corresponding disease or trait outcomes in this sample. For example, the chronic kidney disease (CKD) PRS analysis identified circulating creatinine as by far the most strongly associated biomarker (Beta=0.057 per 1-SD change in CKD PRS, 95% CI=0.051 to 0.063, P=7.79×10^−76^). A strong positive relationship between an elevated genetic burden for CKD and creatinine fits with the known biology, as this biomarker is routinely used to help diagnose CKD in a clinical setting and was even used by the CKD GWAS to define cases (Pattaro *et al*., 2016). Our atlas also includes sex-stratified estimates for PRS weighted by GWAS undertaken in female only (such as breast cancer and age at menarche) and male only (e.g. prostate cancer) populations. Furthermore, we include a bespoke PRS for Alzheimer’s disease excluding variants within the *APOE* gene (as well as 500kbs up and downstream of its encoding region) given the interest in harnessing blood metabolites as biomarkers for this outcome (Proitsi *et al*., 2017) as a means of establishing the extent to which *APOE* is responsible for the associations of the Alzheimer’s disease PRS on blood metabolic biomarkers. Investigating the metabolic profile for the full Alzheimer’s disease PRS (i.e., using variants from across the genome including *APOE*) highlighted 85 associations robust to FDR<5%, in particular types of low and intermediate-density related lipoprotein lipid traits (**Supplementary Table 4**). However, running this analysis using the Alzheimer’s PRS excluding genetic variants at *APOE* found that evidence of association for these biomarkers typically attenuated with no findings surviving FDR<5% (**Supplementary Table 5**). This suggests that variants at this region of the genome, which are recognised to exert their influence on phenotypic traits via horizontally pleiotropic pathways (Ferguson *et al*., 2020), may be responsible for driving the associations robust to FDR corrections in the initial analysis.

### Orienting the direction of effect between putative causal relationships using Mendelian randomization

Along with comparing metabolic signatures for a given PRS, our atlas facilitates hypothesis-free evaluations to inspect all PRS associations for a given metabolic trait. As an example of this, we have undertaken such an analysis based on the associations between all 129 PRS in our atlas with circulating glycoprotein acetyls (GlycA). GlycA is a biomarker of chronic inflammation and has been found to predict various endpoints, including types of cardiovascular disease, cancer, and all-cause mortality (Lawler *et al*., 2016, Connelly *et al*., 2017). Although previous studies of genetically predicted GlycA have been conducted for hypotheses regarding single endpoints (Lord *et al*., 2021), whether or not circulating GlycA has a causal effect on outcomes from across the disease spectrum has yet to be comprehensively investigated. The role of GlycA is important to establish given the emerging role of inflammation as a pharmacologically modifiable pathway for the prevention and treatment of cardiovascular disease.

There were 49 PRS associations with GlycA which were robust to a FDR<5%, used as a heuristic to determine which results to investigate in further detail (**Figure 2 & Supplementary Table 6**). We firstly applied the inverse variance weighted (IVW) MR method to systematically assess whether genetic liability to any of these disease endpoints or complex traits provided evidence of an effect on GlycA levels. In total, 9 of these exposures provided evidence of a genetically predicted effect based at FDR<5% (**Supplementary Table 7**), which included anthropometric traits such as BMI (Beta=0.16, 95% CI=0.11 to 0.21, P=9.9×10^−10^) and genetic liability to cigarettes smoked per day (Beta=0.28, 95% CI=0.20 to 0.35, P=2.4×10^−14^). Estimates based on the IVW method were typically supported by the weighted median approach, although only cigarettes smoked per day were supported by both the weighed median (Beta=0.25, 95% CI=0.16 to 0.34, P=5.5×10^−8^) and MR-Egger (Beta=0.23, 95% CI=0.08 to 0.37, P=0.02) methods (**Supplementary Table 8**).

**Figure 2:**
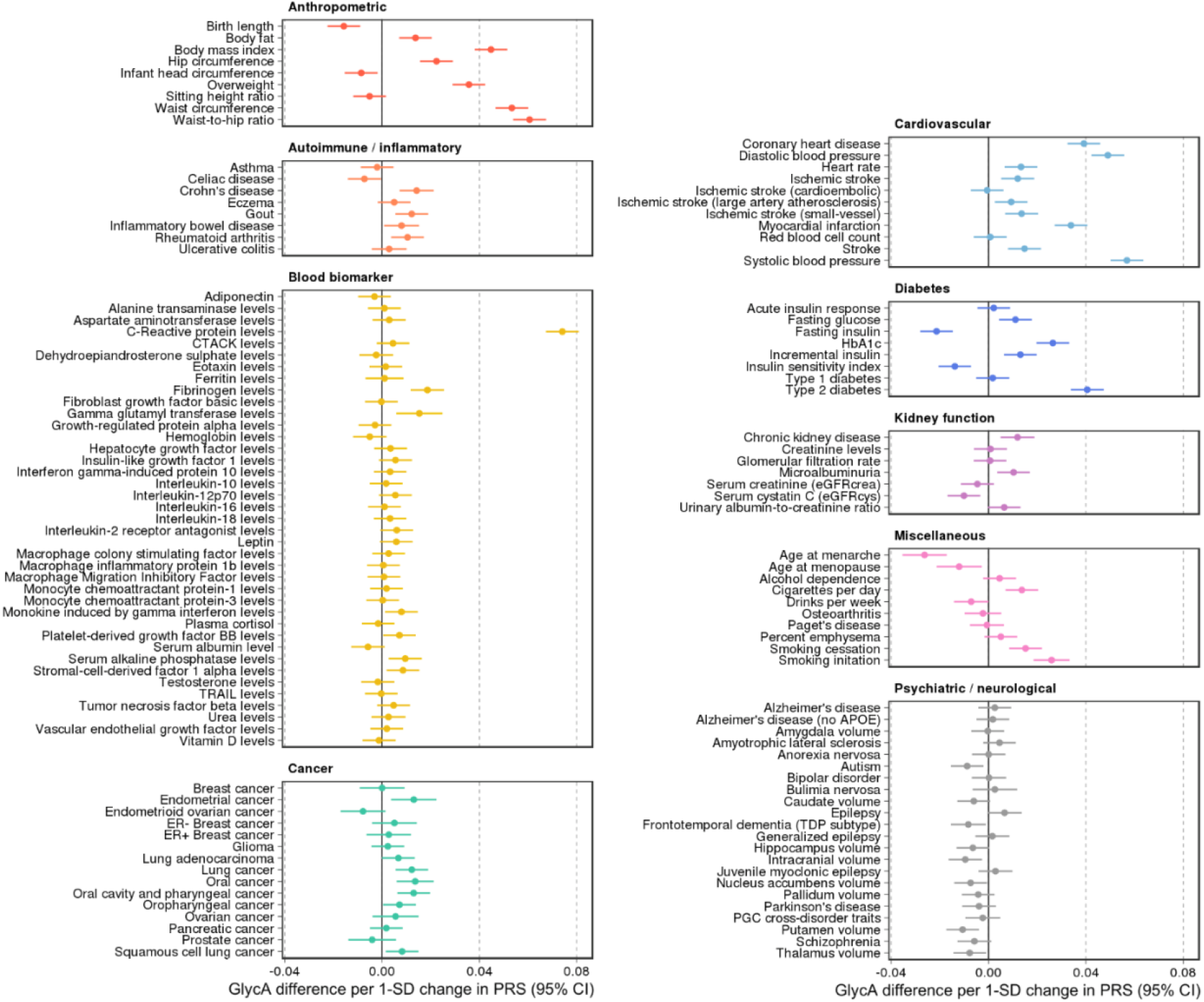
Forest plots depicting results from a systematic evaluation of 129 polygenic risk scores and their associations with circulating glycoprotein acetyls (GlycA).

Next, we investigated the converse direction of effect using MR to assess whether genetically predicted GlycA may influence any of the 49 complex traits or disease endpoints highlighted by our atlas of results. Undertaking a GWAS of GlycA in the UKB identified 59 independent genetic variants which were harnessed as instrumental variables (mean F=100.1) (**Supplementary Table 9**). In contrast to the previous analysis, we identified very weak evidence using the IVW method that genetically predicted GlycA has an effect on any of the traits or diseases assessed based on FDR<5% (**Supplementary Table 10**).

### Stratifying analyses by age to investigate potential sources of bias induced by medication use

A critical challenge when analysing the NMR metabolites data in UKB concerns the most appropriate manner to account for participants taking medications which may undermine inference (Bell *et al*., 2021). For example, UKB participants undergoing statin therapy will likely have altered levels of lipoprotein lipid metabolites compared to others. However, adjusting for statin therapy as a covariate or by stratification can induce collider bias, which may be encountered when investigating the relationship between two factors (such genetic liability towards coronary heart disease (CHD) and a lipoprotein lipid metabolite) which both influencing a third factor (e.g., statin therapy) (**Supplementary Figure 3**). In particular due to the large sample sizes provided, collider bias in the UKB study has been shown to distort findings (Griffith *et al*., 2020) and in extreme cases can even result in opposite conclusions being drawn (Richardson *et al*., 2019a). Therefore, to investigate the influence of medication use on the results within our atlas, we repeated all analyses stratified by age tertiles as proposed previously (Bell *et al*., 2021), given that age is very unlikely to act as a collider between PRS and circulating metabolites, and medication use is likely to be lower in the younger tertiles. Comparisons between the youngest and oldest tertiles in UKB can be systematically investigated and visualized using our web application to evaluate how medications may bias findings.

**Figure 3:**
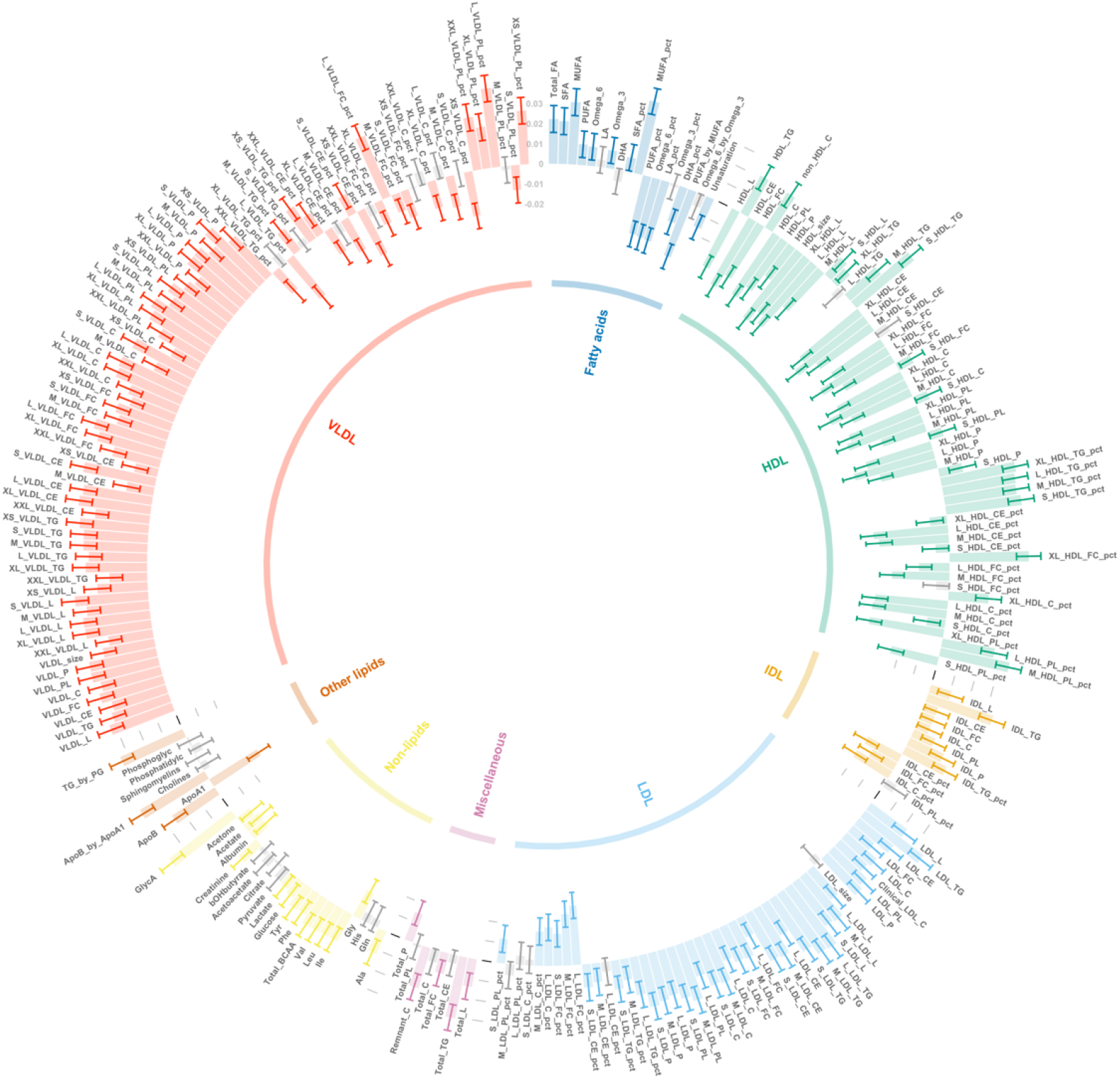
A circos plot illustrating the associations between the polygenic risk score (PRS) for coronary heart disease (CHD) (P<0.05) with 249 circulating metabolic traits in UK Biobank. Error bars represent confidence intervals of the effect estimates and are oriented such that those extending to the outer rim reflect a positive association between the CHD PRS and metabolic traits whereas those extending inwards indicate inverse associations. Associations are coloured and clustered based on subclasses of metabolic traits: between the CHD PRS (P<0.05) and traits from the 6 subclasses: HDL = high density lipoprotein lipids, IDL = intermediate density lipoprotein lipids, LDL = low density lipoprotein lipids, VLDL = very low density lipoprotein lipids. Grey bars represent association with false discovery rates (FDR)>5%.

As an example of this, in the full UKB sample there were 214 circulating metabolites associated with the PRS for CHD (based on the P<0.05 & r^2^<0.1 criteria) based on FDR<5% (**Supplementary Table 11**). The vast majority of these were lipoprotein lipid traits, which are well established causal risk factors for CHD risk. A circos plot depicting all CHD PRS associations with traits which capture lipoprotein lipid concentrations on the NMR panel can be found in **Figure 3**. Amongst the top associations for this PRS was apolipoprotein B (apo B) (Beta=0.025, 95% CI=0.019 to 0.032, P<1×10^−300^), which acts as an index of the number of circulating atherogenic lipoprotein particles and has been postulated previously to be the predominating lipoprotein lipid trait which drives CHD risk (Ference *et al*., 2019, Sniderman *et al*., 2019, Richardson *et al*., 2020b, Richardson *et al*., 2021)

Evaluating this association between age tertiles allowed us to investigate whether it may be influenced by medications in UKB, such as the impact of statin therapy on lowering LDL cholesterol, which apo B particles carry. In the youngest tertile (mean age=47.3 years, 5% statin users), the association between the CHD PRS with apo B was markedly stronger than in the total sample (Beta=0.059, 95% CI=0.048 to 0.069, P<1×10^−300^). In stark contrast, there was weak evidence of an association between apo B and the CHD PRS in the oldest tertile (mean age=65.3 years, 29% statin users) (Beta=-0.009, 95% CI=-0.021 to 0.003, P=0.141), which is likely attributed to the higher proportion of participants undergoing statin therapy in this sample. Similarly, concentrations of VLDL, LDL and IDL provided evidence of a positive association with the CHD PRS in the youngest tertile (**Figure 4a**), whereas the corresponding associations in the oldest tertile provided weak evidence of association (and in some cases reversed direction entirely) (**Figure 4b**). A comparison of all 249 associations with the CHD PRS derived in the youngest and oldest age tertiles can be found in **Supplementary Table 12**, where a large degree of heterogeneity was identified between the youngest and oldest age strata (Q=3618.6, P<1×10^−300^).

**Figure 4:**
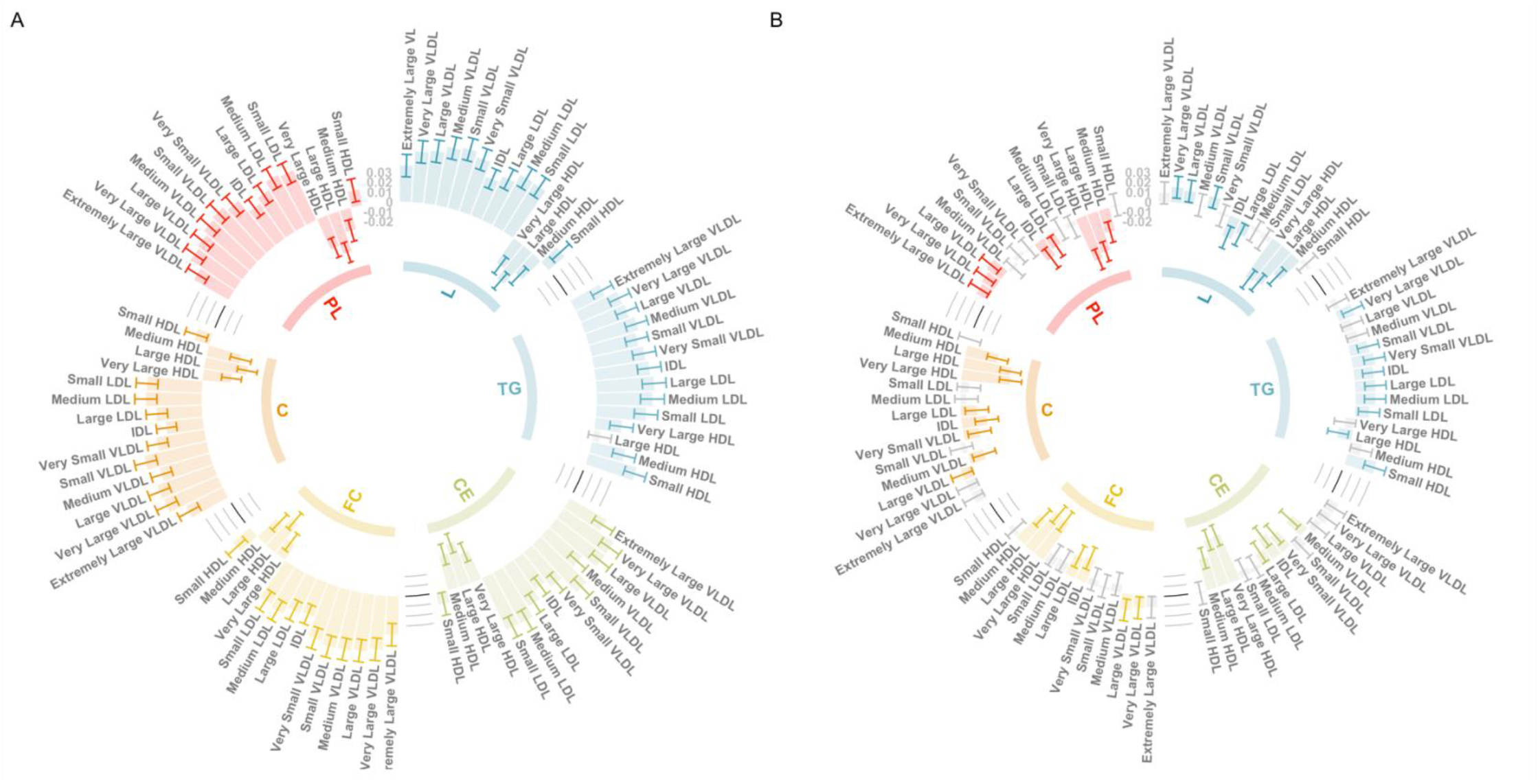
Circos plots illustrating the utility of age-stratified analyses in UK Biobank to investigate potential sources of bias when evaluating associations between polygenic risk scores (PRS) and circulating metabolites. Error bars represent confidence intervals of the effect estimates between the coronary heart disease (CHD) PRS (P<0.05) and traits from the 6 subclasses: L=total lipids, TG=triglycerides, CE=Cholesteryl esters, FC=free cholesterol, C=cholesterol, PL=phospholipids. Grey bars represent association with false discovery rates (FDR)>5%. These barcharts are oriented such that those extending to the outer rim reflect a positive association between the CHD PRS and metabolic traits whereas those extending inwards indicate inverse associations. A) Analyses undertaken for participants in the lowest age tertile (mean age=47.3 years, 5% statin users) and B) The corresponding results for the oldest age tertile (mean age=65.3 years, 29% statin users).

## Discussion

In this study, we have developed an atlas of polygenic risk scores associations with circulating metabolic traits in an unprecedented sample size compared to previous studies. Our results can be used to help prioritise findings worthy of follow-up, using techniques such as Mendelian randomization, as a means of disentangling putative causal and non-causal relationships underlying associations between PRS and circulating biomarkers. Furthermore, conducting all analyses within age tertiles illustrates the potential of medication use within the UKB population to bias relationships within our atlas of results. These results should help highlight disease-metabolic trait relationships where researchers should exercise caution when interpreting findings from their own analyses of the recently generated NMR metabolites data in UKB, which are due to be available in all ∼500,000 participants in the forthcoming years.

Amongst the thousands of PRS associations identified in this study with an FDR<5%, we observed an enrichment of scores derived using GWAS of anthropometric traits. This was exemplified by the body mass index (BMI) PRS which yielded the largest number of associations in our atlas (n=226). Previous studies in the field have demonstrated the strong influence that adiposity has on circulating traits from across the metabolome (Wurtz *et al*., 2014), and indeed across the proteome (Folkersen *et al*., 2020). Furthermore, as shown previously by a Mendelian randomization study (Bell *et al*., 2021), certain associations with the BMI PRS may be due to the influence of medication use in the UKB sample, for example those related to low density lipoprotein (LDL) (e.g. total lipids in LDL: Beta=-0.022, 95% CI=-0.029 to -0.015, P<1×10^−300^). In our atlas, evidence of these associations strongly attenuated in the youngest age tertile, where the influence of such factors in the UKB population may be weakest (e.g. total lipids in LDL from youngest age tertile: Beta=2.8×10^−4^, 95% CI=-0.011 to 0.011, P=0.96). In addition to the striking difference highlighted in our study between the CHD PRS and apolipoprotein B between age tertiles, findings such as this further emphasise the importance of evaluating results from the full sample analysis using those derived in age-stratified subsamples.

As an exemplar, we conducted a hypothesis-free evaluation of one of the metabolic traits on the UKB NMR panel, glycoprotein acetyls (GlycA), as a means of demonstrating how findings from our atlas may help generate hypotheses and follow-up analyses. Whilst our MR results indicated that modifiable risk factors such as BMI and cigarette smoking may increase levels of this circulating inflammatory biomarker, they suggest that targeting GlycA itself is unlikely to yield a beneficial therapeutic effect on the complex traits and disease endpoints evaluated in this study. This highlights the value of findings from our atlas, complemented by approaches such as MR, to help both prioritise and deprioritise circulating metabolic traits for further evaluation. Similar hypothesis-free evaluations on the other 248 metabolic traits can be routinely undertaken using our web tool, in addition to evaluations using the more stringent PRS construction criteria of P<5×10^−8^ and r^2^<0.001. We reiterate the importance of using approaches such as MR (including sensitivity analyses, which are at least partially robust to various forms of pleiotropy) to formally assess putative causal relationships which may underlie findings in our atlas however, as well as to help orient their directionality. This is particularly important given that PRS may be more prone to recapitulating sources of bias commonly encountered in observational studies in comparison to formal MR analyses (Richardson *et al*., 2019b).

In terms of study limitations, we note that the NMR panel is predominantly focused on lipoprotein lipids and as such our atlas does not facilitate analyses across the entire metabolome. Availability of metabolomics quantified by other platforms (e.g. mass spectroscopy) in large numbers with GWAS genotyping will aid in this effort (Lotta *et al*., 2021). Furthermore, whilst these data provide an unparalleled sample size compared to predecessors, findings are based on traits derived from whole blood and may therefore not be reflective of molecular signatures identified in other tissue types (Richardson *et al*., 2020a). In terms of interpretation, we emphasise that PRS can capture an estimate of an individual’s lifelong disease risk, and as such results based on the UKB NMR metabolites dataset, measured at a midlife timepoint in the lifecourse in predominantly healthy participants, may differ substantially to metabolomic profiles of patients with a disease. Conversely, findings may hold the potential to highlight biomarkers useful for disease prediction before clinical manifestation, therefore indicating a potential window of opportunity for early detection and/or intervention. Lastly, in this study we leveraged data from the European subset of the UKB study, which may therefore not be representative of individuals from other ancestries (Duncan *et al*., 2019). Larger sample sizes of non-European individuals with metabolomics data will facilitate analyses in other ancestries once available, in addition to findings from future large-scale GWAS which have been principally confined to individuals of European descent to date (Sirugo *et al*., 2019).

We envisage that findings from our atlas will motivate future study hypotheses and help prioritise (and deprioritise) circulating metabolic traits for further in-depth research. Although we highlight several key findings in this manuscript, all our findings can be queried using our web application which provides a platform to inform researchers in the field planning similar analyses. Similar evaluations to those conducted in this manuscript should help develop a deeper understanding into how circulating metabolic traits contribute towards complex trait variation and assess their putative mediatory roles along the causal pathways between modifiable lifestyle risk factors and disease endpoints.

## Methods

### Data sources

#### The UK Biobank study

Metabolic profiling was undertaken on a random subset of individuals from the UK Biobank (UKB) study (Sudlow *et al*., 2015) (range between 116,353 to 121,695). Details on genotyping quality control, phasing and imputation in UKB have been described previously (Bycroft *et al*., 2018). In total, 249 metabolic biomarkers were generated using non-fasting plasma samples (aliquot 3) taken from UKB participants at initial or subsequent clinical visits. Targeted high-throughput NMR metabolomics from Nightingale Health Ltd (biomarker quantification version 2020) were used to generate data on each of the 249 measures. These included biomarkers on lipoprotein lipid traits, their concentrations and subclasses, fatty acids, ketone bodies, glycolysis metabolites and amino acids. Further details are described elsewhere (Julkunen *et al*., 2021). Statin users in UKB were identified based on medication codes as defined previously (Sinnott-Armstrong *et al*., 2021). A full list of these metabolic biomarkers and their summary characteristics can be found in **Supplementary Table 1**.

Ethical approval for this study was obtained from the Research Ethics Committee (REC; approval number: 11/NW/0382) and informed consent was collected from all participants enrolled in UKB. Data was accessed under UKB application #15825.

#### Genome-wide association study summary statistics

Publicly available GWAS summary statistics were extracted from the OpenGWAS platform (https://gwas.mrcieu.ac.uk/) and publicly available repositories (Elsworth *et al*., 2020). We identified GWAS for 129 different complex traits and diseases which were selected to encompass a broad range of human phenotypes for which genome-wide data was available allowing us to construct PRS based on all variants with P<0.05. Furthermore, we identified GWAS based on study populations with participants of European descent, as our study was based on the unrelated European participants of UKB with NMR measures, as well as studies which had not analysed the UKB study population to avoid overlapping samples which can lead to overfitting bias in results (Fang *et al*., 2021). All detail of these GWAS can be found in **Supplementary Table 2**. The only exceptions to this were systolic and diastolic blood pressure which were based on a GWAS meta-analysis which included UKB (Evangelou *et al*., 2018), as no UKB-independent GWAS of European individuals for these traits was available from OpenGWAS.

#### Polygenic risk score construction

We built two versions of each PRS in this study using the following criteria. Firstly, scores were developed with independent variants (i.e. r^2^<0.001) which were robustly associated with their traits or disease based on conventional genome-wide corrections (i.e. P<5×10^−8^). The second versions of scores were derived using more lenient thresholds which were r^2^<0.1 and P<0.05. Linkage disequilibrium (LD) to estimate correlation between variants was based on a previously constructed reference panel of 10,000 randomly selected unrelated UKB individuals of European descent (Kibinge *et al*., 2020). PRS were derived for all participants with both genotype and NMR metabolites data after firstly excluding individuals with withdrawn consent, evidence of genetic relatedness or who were not of ‘white European ancestry’ based on a K-means clustering (K=4). These scores were built by summing trait/disease risk increasing alleles which participants harboured weighted by their effect size reported by GWAS using the software PLINK (Chang *et al*., 2015).

The majority of PRS were constructed in all eligible participants, with the exception of those based on GWAS in sex-stratified populations. These were breast cancer (including ER+ and ER-PRS), endometrial cancer, ovarian cancer, endometrioid ovarian cancer, age at menarche, age at menopause and bulimia nervosa, which were derived in females only, as well as the prostate cancer PRS derived in males only. We also built a bespoke PRS for Alzheimer’s disease excluding variants at the *APOE* gene locus (consisting of the gene region itself as well as a 500kb window either side) in all eligible participants given the known pleiotropic role in lipid metabolism (Ferguson *et al*., 2020).

### Statistical analysis

#### Polygenic risk score analysis

To allow us to draw comparisons between PRS-metabolite associations, we standardized all PRS to have a mean of 0 and standard deviation of 1 and additionally applied inverse rank normalization transformations to all metabolic traits prior to analysis. Associations between PRS and normalized metabolites were determined by linear regression with adjustment for age, sex (where appropriate) and the top 10 principal components. All analyses were undertaken using both versions of each PRS as long as their corresponding GWAS had at least one variant with P<5×10^−8^ necessary for the more stringent criteria. Each analysis was conducted initially in the full sample, followed by analyses after stratification into age tertiles to investigate the influence of medication use on findings. To account for multiple testing in this study, we applied a Benjamini-Hochberg false discovery rate threshold (FDR) of less than 5% as a heuristic to highlight finding worthwhile evaluating in further detail. However, all results from our analyses are available in the web application.

#### Instrument selection for glycoprotein acetyls analysis

Genetic instruments for all PRS traits/disease points evaluated in this study using MR were obtained from the ‘TwoSampleMR’ v0.5.6 R package (Hemani *et al*., 2018), or by manually uploading GWAS summary statistics and using the ‘clump_data’ function from this package to identify them. Instruments for glycoprotein acetyls (GlycA) were identified by conducting a GWAS of this trait in the UK Biobank study using the BOLT-LMM (linear mixed model) software to control for population structure (Loh *et al*., 2015). Analyses were undertaken after excluding individuals of non-European descent (based on K-mean clustering of K=4) and standard exclusions, including withdrawn consent, mismatch between genetic and reported sex, and putative sex chromosome aneuploidy. Analyses were adjusted for age, sex, fasting status and a binary variable denoting the genotyping chip used in individuals (the UKBB Axiom array or the UK BiLEVE array). Genetic instruments were defined as variants with P<5×10^−8^ after removing those in linkage disequilibrium using the ‘clump_data’ function as above.

#### Mendelian randomization analyses

MR analyses were undertaken using the ‘TwoSampleMR’ package (Hemani *et al*., 2018) to estimate the bi-directional effects between PRS traits/disease endpoints and GlycA. This was firstly estimated using the inverse variance weighted (IVW) method, which takes the SNP-outcome estimates and regresses them on those for the SNP-exposure associations (Burgess *et al*., 2013), followed by the weighted median and MR-Egger methods which are considered to be more robust to horizontal pleiotropy than the IVW approach (Bowden *et al*., 2015, Bowden *et al*., 2016). F-statistics were calculated to assess weak instrument bias (Burgess and Thompson, 2013).

Forest and circos plots in this study were generated using the R package ‘ggplot’ v3.3.3 (Ginestet, 2011). Heatmaps were generated using the R package ‘pheatmap’ v1.0.12 (Kolde, 2015). The web application was developed using the R package ‘shiny’ v1.0.4.2 (Chang *et al*., 2020). All analyses were undertaken using R (version 3.5.1).

## Data Availability

All data produced in the present work are either contained in the manuscript or available online at http://mrcieu.mrsoftware.org/metabolites_PRS_atlas/.

## Acknowledgements

We would like to thank all the consortia for making their summary statistics available for the benefit of this work. We also thank the participants of the UK Biobank study. Data was accessed under UKB application #15825.

