## Supplementary Figures for "An atlas of associations between polygenic risk scores from across the human phenome and circulating metabolic biomarkers": Supplementary_Figures_legends.docx

**Supplementary Figure Legends**

**Supplementary Figure 1.**

Heatmap showing the Z scores of associations between metabolites and PRS that were derived using the lenient criteria (i.e., variant-trait associations (P) <0.05 and linkage disequilibrium (LD) r^2^<0.1). Each row represents a metabolite, and each column represents a PRS. Metabolites and PRS are grouped by their subcategories, which are annotated on the left and top of the heatmap, respectively. Dark red represents a higher Z score, dark blue represents a lower Z score.

**Supplementary Figure 2.**

Heatmap showing the Z scores of associations between metabolites and PRS that were derived using the more stringent criteria (i.e., variant-trait associations (P) <5x10^-8^ and linkage disequilibrium (LD) r^2^<0.001). Each row represents a metabolite, and each column represents a PRS. Metabolites and PRS are grouped by their subcategories, which are annotated on the left and top of the heatmap, respectively. Dark red represents a higher Z score, dark blue represents a lower Z score.

**Supplementary Figure 3.**

Directed acyclic graphs (DAGs) showing the collider and confounders in the effects from coronary artery disease to metabolites in the UK Biobank.
