## Supplementary figures and images for "An atlas of associations between polygenic risk scores from across the human phenome and circulating metabolic biomarkers"

### Figure S1.png

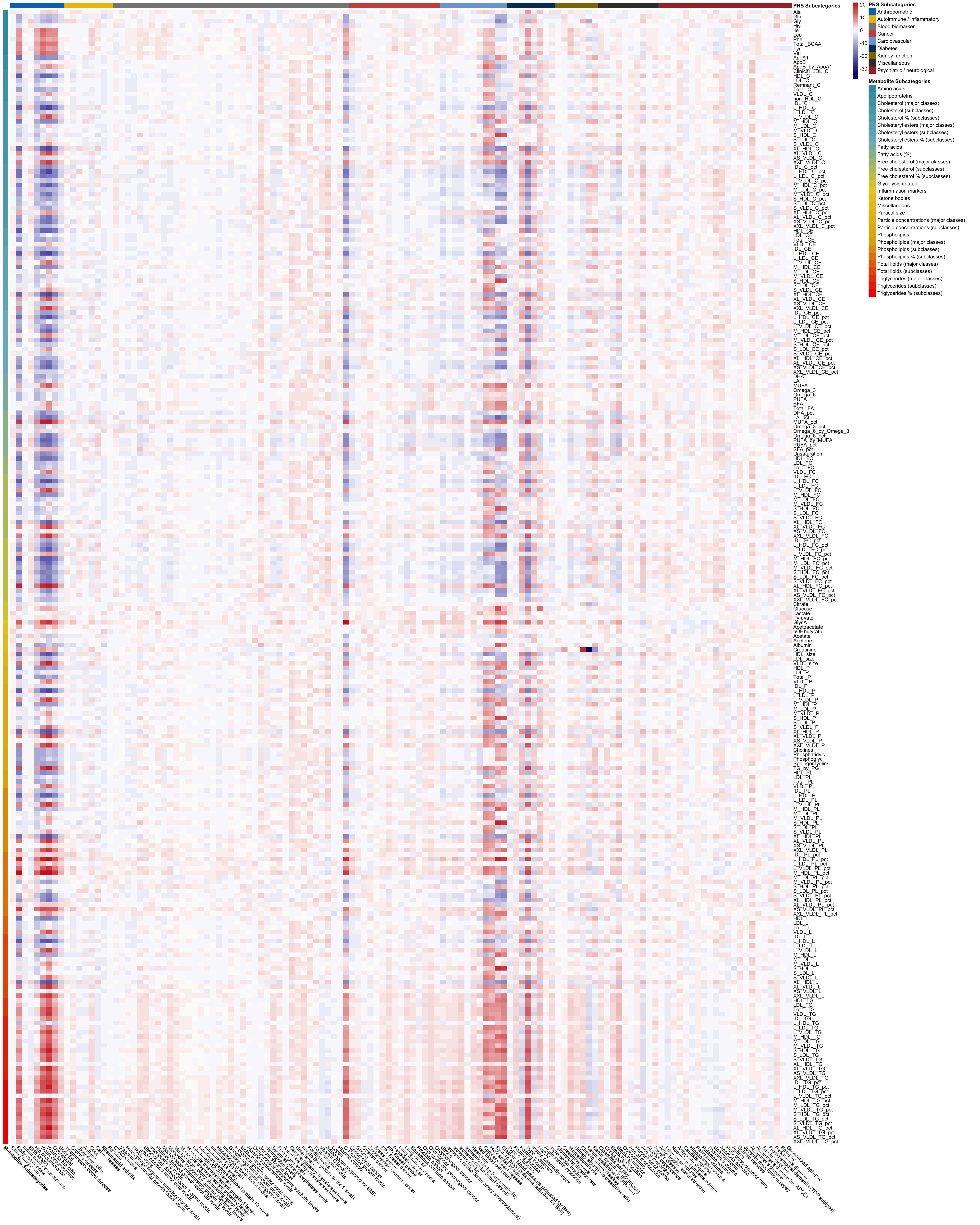

### Figure S2.png

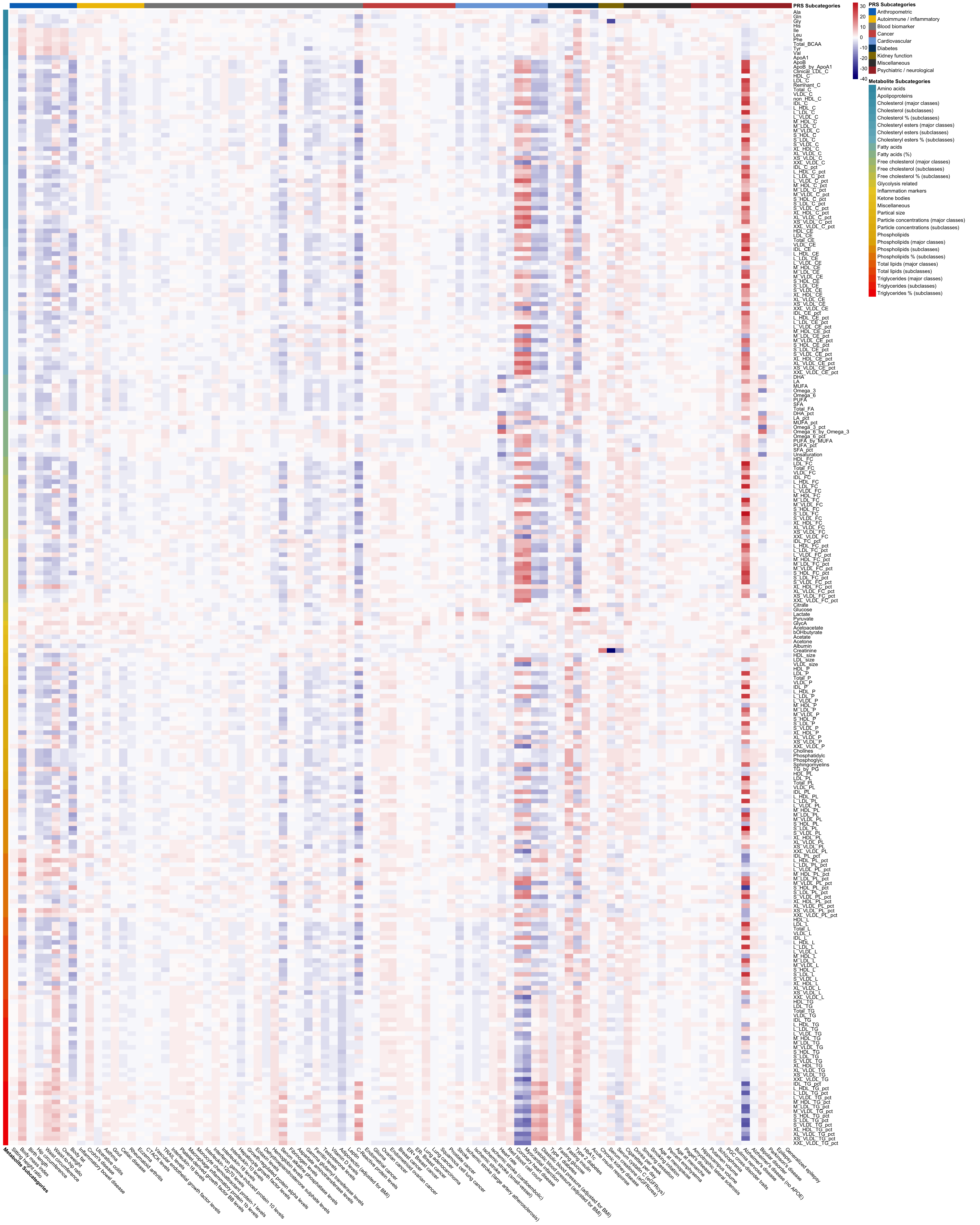

### Figure S3.png

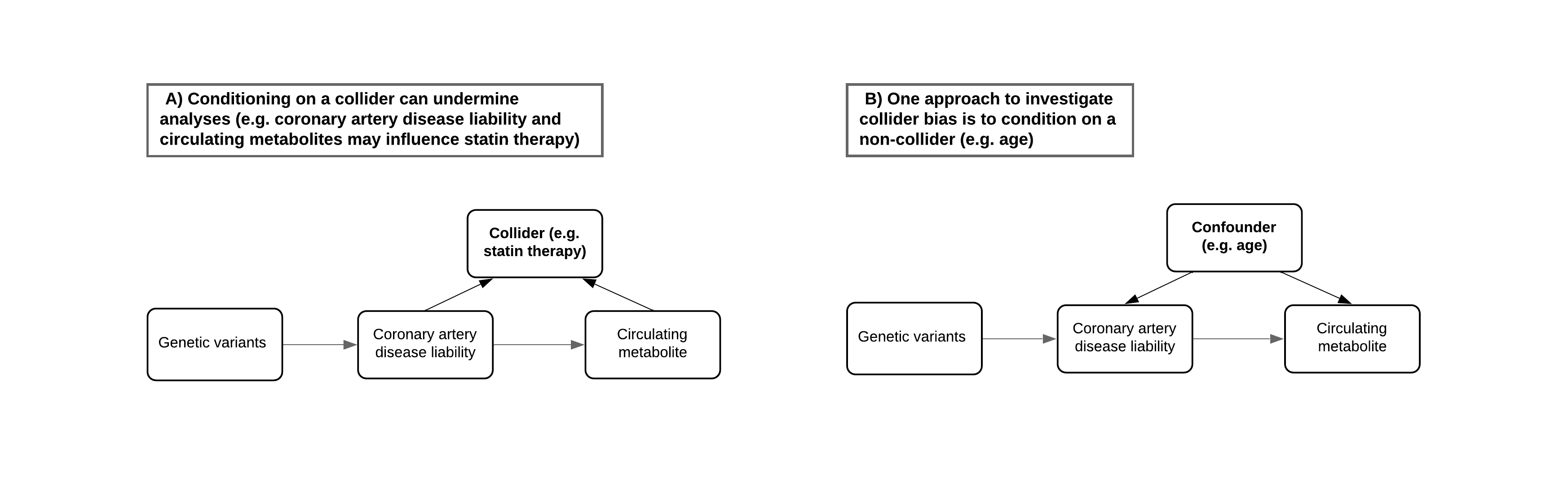
